# Application of Extreme Gradient Boosting to predict NCD-HIV/AIDS comorbidity in young adults in Malawi

**DOI:** 10.1101/2025.08.05.25333013

**Authors:** Ansley Kasambara, Mphatso S. Kamndaya, Salule J. Masangwi, Atupele Mulaga

**Author notes:** Corresponding author: Ansley Kasambara.

## Abstract

The fight to achieve Sustainable Development Goal 3.3 and 3.4 by 2030 requires data driven approaches and appropriate methodologies that would enable sound analyses of data. Non-communicable diseases and the comorbidity of non-communicable diseases and HIV/AIDS are prevalent in Sub-Saharan Africa. However, there is limited data to substantiate the burden in order to arrive at decisions on management and interventions. This study developed a model for prediction of NCD and NCD-HIV/AIDS cases among young adults using a machine learning method.

A retrospective study design in which 17741 patient level data was collected from NCD Mastercards. A sub sample of 2763 young adults was selected and a machine learning algorithm was used to develop multiclass models on NCDs only and NCD-HIV/AIDS. Specifically, Extreme Gradient Boosting was used to model classification of NCD as well as NCD-HIV/AIDS cases.

The NCD and NCD-HIV/AIDS comorbidity models were developed and classified different cases given patients socio-demographic features. The models performed well with both training and validation loss below 0.5 compared to 0.8 threshold. Metrics for the overall goodness of the models were all above 0.8 indicating particularly good model performance. Accuracy of the NCD only and NCD-HIV/AIDS models were 68% and 84%, respectively. The most influential factors among others for prediction in the NCD only and NCD-HIV/AIDS models were having no intervention (26.8%) and living in a city (45.6%) respectively.

The models by the XGBoost algorithm correctly classified non-communicable diseases and NCD-HIV/AIDS comorbidity given a set of socio-demographic factors such as gender, different location aspects and availability of intervention. The models could be deployed and used for basic predictions on suspected individuals given their socio-demographic factors. Furthermore, the socio-demographic factors that significantly influence an individual having a non-communicable disease or NCD-HIV/AIDS would be used to design appropriate interventions to control the increase in the number of new cases.

## Introduction

The increasing trajectory in non-communicable disease cases have been a worldwide concern [1–3]. The developed world has had a slowdown in the number of cases whereas the developing has seen significant growth in number of cases due to the advent of risk factors [1,4]. Sub Saharan Africa region being one of the most affected areas among the developing countries, which includes Malawi [2,3]. Additionally, HIV/AIDS has been an epidemic which has a dreadful impact worldwide since 1981 [5,6]. The number of new HIV infections has been reducing year by year in most countries coupled with antiretroviral therapy which has reduced the number of fatalities [7]. However, sub-Saharan Africa remains largely affected by HIV/AIDS [8,9]. As such an intersection is observed where an individual can be afflicted by both NCDs and HIV /AIDS which entails a deadly comorbidity that needs attention [10–12].

Non-communicable Diseases and HIV/AIDS risk factors are more prevalent in developing countries due to growth in economic activities, rapid urbanisation, and industrialisation [13,14]. The sub-Saharan Region has seen growth in several cities Including Malawi’s own Blantyre, Lilongwe, Mzuzu and Zomba fostered by urbanisation and industrialisation [10,11]. As such it can be observed that location plays a significant role in the growth of NCD and HIV/AIDS cases [15,16]. District locations such as those in lakeshore areas and borders may also be characterized by high economic activities because of tourism and foreign currency exchange respectively - these also propel the existence of NCD and NCD-HIV/AIDS risk factors [14]. Malawi’s landscape has both border and lakeshore districts [17,18]. This puts Malawi in a position susceptible to increased NCD and NCD-HIV/AIDS cases.

The growth in NCDs and HIV/AIDS comorbidity cases is more pronounced among the older age group [19]. This stems from the fact that NCDs are more prevalent among old people [20]. On the contrary, NCDs are now affecting young and young adult individuals, which changes the dynamics of NCD management since a wider age group is affected [10,21]. Furthermore, for people living with HIV, ART enables them to live longer into old age, which puts them at risk of NCDs, hence the existence of NCD-HIV/AIDS comorbidity [22]. However, HIV/AIDS does not discriminant on age hence all age groups are at risk [23–25]. A particular age of concern is the young adult age group, which essentially drives the economy of a country [26,27]. This is the most productive age group, such that days off work are detrimental to their families, organisations, and country as a whole [28,29].

Non-communicable diseases and HIV/AIDS pose a great concern in the health sector, such that the framers of the Sustainable Development Goals specifically under Goal 3, explicitly focus on NCDs and HIV/AIDS under clauses 3.3 and 3.4, respectively [4,30]. These goals provide targets to be attained for each ailment by 2030 [30,31]. Malawi, at the country level, has framed the Malawi 2063 Agenda in which health is one of the priority areas [32]. This health agenda is fed by the National Health research Agenda II, whose focus is also on managing NCDs, HIV/AIDS, and particularly the NCD-HIV/AIDS comorbidity [33]. Research being core to the development of programs and planning interventions, the National Health research Agenda II calls for studies focusing on these thematic areas [33].

The need for data-driven approaches for the management of NCDs and NCD-HIV/AIDS comorbidity has led to the lack of readily available data [7,10,34]. Access to data, even if it is not all the required data, enables modelling that will provide estimates and/ or predictions necessitating quick responses to upward trends [35,36]. World Health Organisation targets for NCDs are below the requirements for Malawi, and data availability being one of the components that the country has not achieved [37,38]. This implies that actionable estimates, forecasts, and predictions cannot be carried out [10]. It is thoroughly lacking to base decisions on NCDs and NCD-HIV/AIDS comorbidity on the premise that cases are increasing worldwide as well as within the region and not within country data [35,36]. Therefore, it is difficult to plan and implement effective interventions that are backed by data.

Further consideration of this study was a specific age group who are at risk of both NCDs and NCD-HIV/AIDS comorbidity but are rarely considered [39,40]. The young adult age group is also affected by these ailments, although the impact is not as large as observed in adults [40,41]. However, young adults have a high potential of contributing to the country’s economy, therefore being afflicted by NCDs and NCD-AIDS comorbidity has a huge impact due to having days off work because of illness or presenteeism [26,42,43]. This has a ripple effect, as in the long run, this affects their productivity at work and subsequently the economy of the country [26,27,43].

The main objective of the study was to develop models to predict NCD and NCD-HIV/AIDS cases among young adults. Several models were considered to achieve this objective. Multinomial models and hierarchical multinomial models, ranging from univariate, multivariable, and multivariate as well as nested models, were conducted [44]. All the models that were employed yielded insignificant results that could not be used to draw conclusions; thus, traditional statistics did not yield significant results and therefore compelled the study to seek machine learning approaches [45,46]. Specifically, Extreme Gradient Boosting (XGBoost), a versatile model was engaged to develop classification models for predicting NCD and NCD-HIV/AIDS comorbidity cases [47,48]. Extreme gradient boosting models have not been employed in Malawi and within the region for such a classification task.

## Methods

A retrospective study whose aim was to predict the categorical response variables, a specific NCD and NCD-HIV/AIDS comorbidity using a set of predictor features, which comprised of socio-demographic factors. Due to the nature of our response variables, the study involved multi-class classification.

Data for this study was extracted from NCD patient Mastercards from 12^th^ September, 2022 to on 19^th^ February, 2023. A Mastercard is a hard copy patient record card that contains the patients’ and the specific ailment details. However, patient identification information was not extracted. The study required data on NCDs and HIV/AIDS and this data was only available from the NCD Mastercard. The data from the mastercards was extracted using an extraction sheet that was deployed using KoboToolBox. Trained research assistants extracted the data using android devices from the selected health facilities. The inclusion criteria for health facilities required that the facility should have been actively using the NCD mastercards. Those facilities that were not using master cards and those that were not in the purposively sampled cities/districts were excluded. The selected cities were Blantyre, Lilongwe, Mzuzu and Zomba whereas districts included Mwanza, Neno, Mangochi, Mchinji, Salima, Nkhatabay and Karonga due to the presence of high-risk factors of having a NCD and HIV/AIDS and/or NCD-HIV/AIDS comorbidity.

The extracted data included (Table 1):

**Table 1:** Data extracted from NCD patient mastercards.

| Feature | Outcome |
| --- | --- |
| NCD only | Asthma, Diabetes, Hypertension, Hypertension-diabetes, Other NCD |
| NCD-HIV/AIDS | Asthma-HIV/AIDS, Diabetes-HIV/AIDS, Hypertension-HIV/AIDS, Hypertension-diabetes-HIV/AIDS, Other NCD-HIV |
| Gender | Male, Female |
| Age | Age of individual |
| City/District | Blantyre, Zomba, Lilongwe, Mzuzu, Mwanza, Neno, Mangochi, Salima, Mchinji, Nkhatabay, Karonga |
| Type of health facility | Health centre, Hospital, District hospital, Central hospital |
| NCD present | Yes, No |
| Type of NCD | Asthma, Diabetes, Epilepsy, Hypertension, Hypertension-diabetes, and Other NCDs |
| NCD diagnosis date | Date |
| HIV/AIDS present | Yes, No |
| HIV diagnosis date | Date |
| NCD intervention | Yes, No |
| Type of NCD intervention | Medication, Outreach/sensitisation, Physical fitness, None |
| NCD-HIV/AIDS intervention | Yes, No |
| Type of NCD-HIV/AIDS intervention | Medication, Physical fitness, None |
| NCD | Asthma, Diabetes, Hypertension, Epilepsy, Hypertension-diabetes etc |
| NCD-HIV/AIDS Comorbidity | Yes, No |

The initial amount of data extracted had 30,686 patient records. Data pre-processing involved management of inconsistencies, missing values and organizing the data. All patient records that had a missing date feature accounted for 6.4% of the data. A date imputation was possible, but the majority dates fell within the range July 2019 to December 2019. Patient records that were within this time frame, were also removed from the dataset because they comprised pre-July 2019 patient cases that were not newly diagnosed. The final sample after data pre-processing was 17,741 patient records.

Feature engineering led to development of new features from existing ones (Table 2). Based on the NCD feature and HIV/AIDS present, a new feature depicting both NCD only and NCD-HV/AIDS comorbidity specific cases were derived. In addition, the district feature enabled derivation of additional features such as region, city/district location and city district designation. Age groups were derived from the age feature with categories implying young, young adult and adults. It was determined in a previous study that there were about 50 different types of NCDs observed [49] and therefore a new feature labelled top 5 NCD and top 5 NCD-HV/AIDS were created. This feature enabled a palatable data analysis process and interpretable outcomes. To further enable a smooth data analysis, all the predictor features were one-hot encoded since they were all categorical features.

**Table 2:** Feature engineering from existing features.

| Feature | Outcome |
| --- | --- |
| NCD only | Asthma, Diabetes, Hypertension, Hypertension-diabetes, Other NCD |
| NCD-HIV/AIDS | Asthma-HIV/AIDS, Diabetes-HIV/AIDS, Hypertension-HIV/AIDS, Hypertension-diabetes-HIV/AIDS, Other NCD-HIV |
| Age group | 19 - 39 |
| CD location | Border, Inland, Lakeshore |
| CD classification | City, District |
| Region | North, Central, South |
| Comorbidity | NCD Only, Comorbidity |
| Top 5 NCDs or top 5 NCD-HIV/AIDS | Asthma, Diabetes, Hypertension, Hypertension-diabetes, Other NCD, Asthma-HIV/AIDS, Diabetes-HIV/AIDS, Hypertension-HIV/AIDS, Hypertension-diabetes-HIV/AIDS, Other NCD-HIV |

The categorical response variables: NCD only and NCD-HIV/AIDS comorbidity were modelled and predicted using the following predictor features; gender, Age group, region, district, Health facility, city/district location, city/district designation, presence of NCD intervention, type of NCD interventions, presence of comorbidity interventions and type comorbidity intervention.

The study was specifically interested in studying young adults who according to McLoed (2025) is a group of individuals aged between 19 and 39 years old. the study adopted the definition by McLoed despite that there are various definitions of a young adult [50]. The young adult age group was chosen because these individuals are the drivers of economies [27,43]. Young adults are the backbone of development since they are in their prime for productivity [26,43]. It therefore implies that if this age group is afflicted with NCDs or NCD-HIV/AIDS comorbidity, productivity would be affected. As such the study concentrated on modelling and predicting NCD cases and NCD-HIV/AIDS cases among young adults. Therefore, a subset of young adults’ patient data totalling to 2763 was taken from the 17,741 patient records.

Data on NCD cases only and NCD-HIV/AIDS comorbidity for the young adults were subset to two different datasets and each was then divided into training and testing set follow in the 80/20 split ratio [51]. This enabled training the models on 80% of the data then assessing how well the model performs using the 20% of the data from the testing set [51,52]. To ensure validity and reproducibility of the results, a five-fold cross validation was carried out on the training data [53,54]. Cross validation further necessitated model evaluation [55,56].

### Data summaries

Data summaries were conducted through determination of average age and the proportion of gender from the total sampled patients. The proportion of gender for young adults was determined using frequencies segregated by age group. These were presented in a table using proportions and confidence intervals. The demographic features: gender, region, classification of location, designation of location were summarised using frequencies, proportions, and confidence intervals aggregated for NCD only and NCD-HIV/AIDS.

Additional summaries were also conducted through determining frequencies for each of the top 5 NCD and NCD-HIV/AIDS comorbidity patients for every year for which data was collected. The output was presented in a table showing frequencies and proportions.

A chi-square test was employed to assess the association between specific NCDs and whether an individual has HIV/AIDS or not. The outcome was plotted on a bar graph to visualise the association. Further tests of association were conducted between the predictors and the response features, having a specific NCD or NCD-HIV/AIDS comorbidity to determine which predictors significantly influence the response features.

### Model selection

A scalable and efficient machine learning method, extreme gradient boosting (XGBoost), was selected to model the classification of NCDs and NCD-HIV/AIDS comorbidity. XGBoost was versatile in handling features and tuning of hyper-parameters which renders it a more probable ML candidate for this classification task [47].

The multi class classification objective function in XGBoost aims to minimize the loss during training in the prediction accuracy while preventing overfitting with a regularisation term. Since the outcome features had more than two outcomes, the multi:softmax function was used to achieve the classification. The objective function, which is the sum of loss function and the regularisation term, is defined as follows:

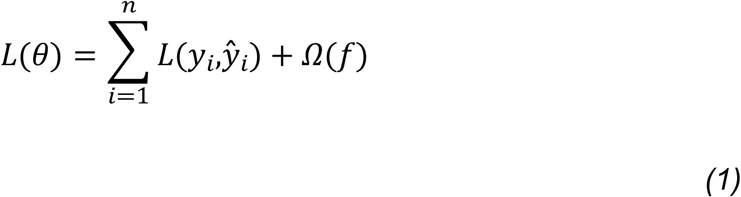

Where:

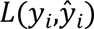 is the loss function for the *i*-th sample, comparing the predicted probability of a specific NCD or NCD-HIV/AIDS comorbidity 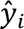, to true class label for NCD or NCD-HIV/AIDS comorbidity *y*_*i*_, for the *i*-th sample and Ω(*f*) is the regularisation term for handling model complexity and prevention of overfitting defined by 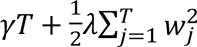.

The softmax function that is used for multi-class classification, calculates the probability distribution over the range of classes for NCD only and NCD-HIV/AIDS comorbidity.

For the number classes for NCD only and NCD-HIV/AIDS comorbidity, denoted by *K*, predicted probability *y*_*ik*_ for a particular class *k* for the *i*-th instance was calculated by

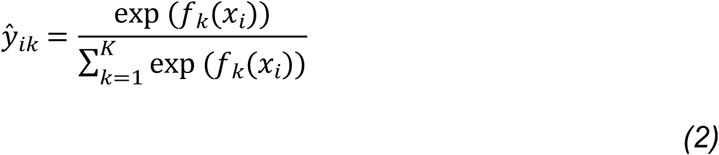

Where:

*f*_*k*_(*x*_*i*_) is the output value for a specific class *k* from the model

The denominator is the total of the exponentials of the output values for all the classes such that the predicted probability adds up to 1.

The cross-entropy loss is the Softmax loss function for the multi-class classification, for a particular class *k* in the *i*-th instance. Cross-entropy loss is defined by

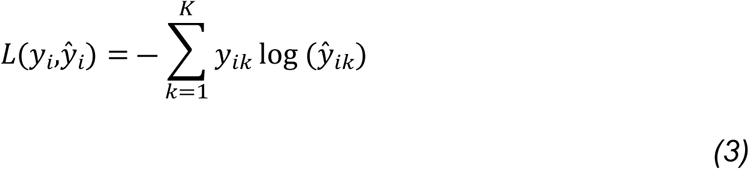

Where:

*y*_*ik*_ is the true label for class *k* (where *k*’s label stands for either, hypertension, diabetes or other NCD for the NCD only model and hypertension-HIV/AIDS, diabetes-HIV/AIDS or other NCD-HIV/AIDS for the NCD-HIV/AIDS model), which is typically one-hot encoded, *y*_*ik*_ = 1 for the correct class and *y*_*ik*_ = 0 for all other classes.

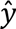_*ik*_ is the predicted probability for specific class *k* for the *i*-th instance.

The XGBoost model is updated sequentially by adding new trees/learners which enables the correction of residual errors of the model.

### Hyperparameter tuning

Extreme gradient boosting has a number of hyperparameters to enable optimization of the classification task to achieve the best model performance [57,58]. Table 3 shows the list of major parameters than can be tuned, their default values, tuning range and values that optimized the best models for NCDs and NCD-HIV/AIDS comorbidity.

**Table 3:** Hyperparameters values for the models.

| Hyperparameter | Description | Default values | Tuning range | NCD Model | NCD-HIV/AIDS Model |
| --- | --- | --- | --- | --- | --- |
| Learning rate (eta) | Determines the contribution of each tree to the mode, | 0.3 | [0,1] | 0.01 | 0.1 |
| Gamma (gamma) | The partitioning of a lead node requires the minimum loss reduction | 0 | $[0, \infty]$ | 0 | 0 |
| Maximum depth (max_depth) | The depth of each tree which determines the complexity of the relationships | 6 | $[0, \infty]$ | 5 | 7 |
| Subsample (subsample) | The proportion of training data that selected to be included in the sample for a given boosting round | 1 | $[0,1]$ | 0.7 | 0.7 |
| Colsample_tree (colsample_bytree) | The proportion of features a selected randomly for a given boosting round | 1 | $(0,1]$ | 0.9 | 1 |
| L2 Regularisation parameter (lambda) | This is used to prevent overfitting hence adds a layer of control to the leaf weights | 1 | $[0, \infty]$ | 1 | 1 |
| L1 Regularisation parameter (alpha) | This is also used to prevent overfitting hence adds a layer of control to the leaf weights | 0 | $[0, \infty]$ | 0 | 5 |

A grid search technique and cross-validation were used to optimise the models hyperparameters which gave the best model performance.

### Model training

The models on classifying NCD only and NCD-HIV/AIDS comorbidity were trained using the training data. The classification was modelled using the chosen hyperparameters. Specifically, the regularisation terms were used to control overfitting and ensuring model generalisation. The model training and validation loss led to unsatisfactory results that would not enable significant prediction. This led to reviewing the predictor features and removing the city/district features from the model. The values of the outcome features were also reduced, then assessing the training and evaluation loss at each level. The models were therefore run as shown in Fig 1 reducing the values of the outcome features and removing the district feature for both models.

**Fig 1.**
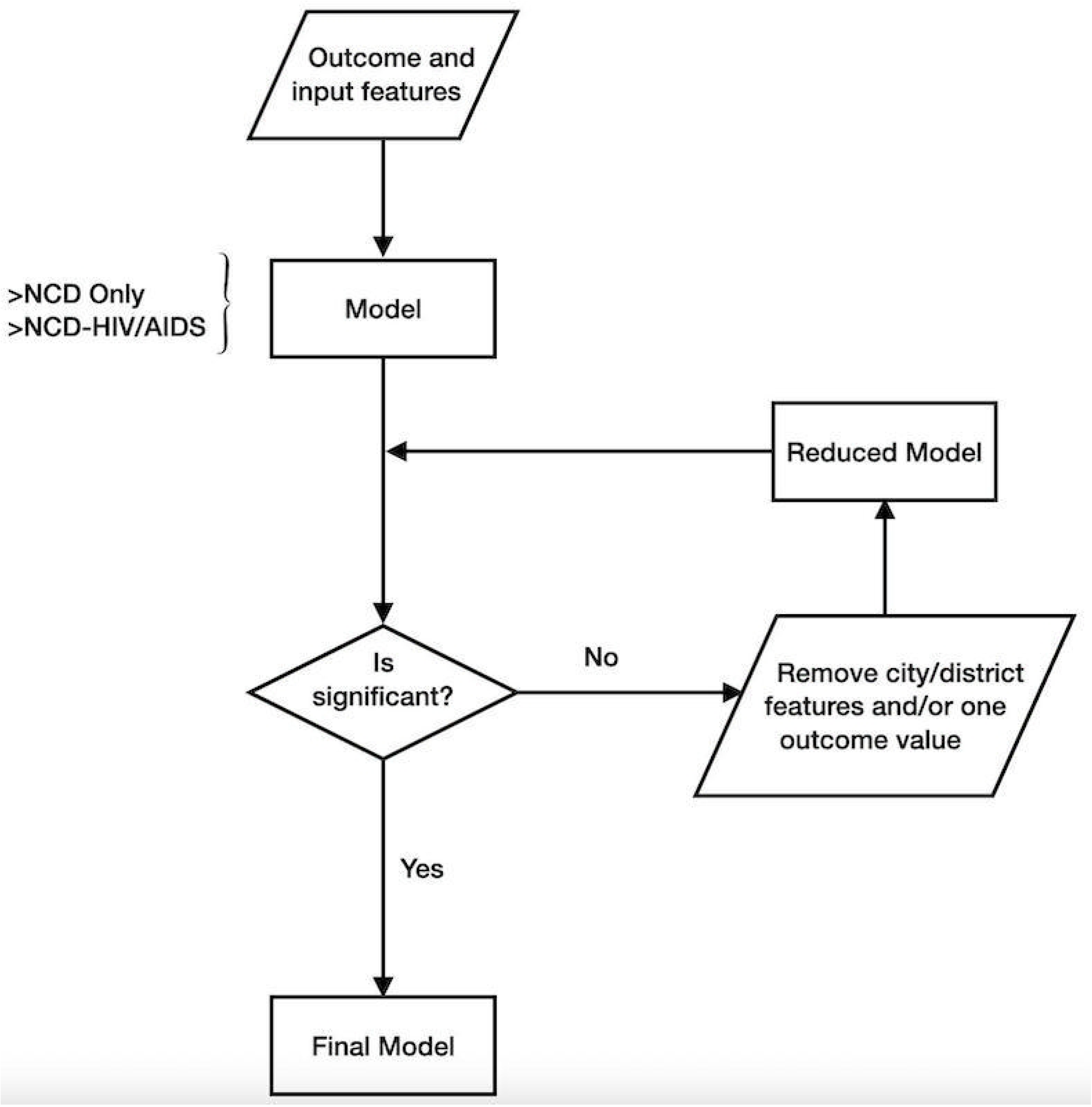
Modelling process to attain the final model adapted from [59].

The model is run with all values of the outcome feature and all the input features. Upon consideration of the training and validation loss, if the loss is above the 0.8 threshold, then the model results are merely by chance and cannot be used for prediction. Furthermore, the difference in the training and validation loss should be as small as possible to ensure that the model performs well on unseen data. In such scenarios, the city/district feature is removed first then run the model again. If the model is still not significant then one of the values of the outcome/response feature is collapsed to “Other NCD.” The cycle continues until a significant model with training and validation loss below the threshold is attained. Both the NCD Only and NCD-HIV/AIDS comorbidity models were significant with no city/district features included and having three values in the outcome/response feature; hypertension, diabetes, other NCD and hypertension-HIV/AIDS, diabetes-HIV/AIDS, other NCD-HIV/AIDS respectively (See appendix 1/additional information).

### Model evaluation

Various performance metrics were utilised to evaluate the models on the test data. The accuracy, precision, recall, macro/F1-weighted scores, receiver operating characteristic (ROC) curve and area under the ROC (AUC-ROC) and the confusion matrix were the chosen evaluation methods for the NCD only and NCD-HIV/AIDS comorbidity models [60–63]. All the evaluation metrics have a range from 0 to 1, where 1 implies a perfect model and 0 a completely wrong model [47,61,62]. The precision, recall, macro/F1-scores have their balance and optimal range between 0.4 and 0.6 with better predictions when metrics are above 0.6 (60%)[64–66].

### Model interpretation

There are number of tools that can be used to interpret XGBoost models. This study employs feature importance tool to draw interpretations from the models [62,67]. The predictions were determined by the contribution of each feature to the model by considering feature importance [59]. This is summarised and displayed using a bar chart.

## Results

There was a total of 17741 patients of which 2763 were young adult patients. Table 4 summarises the distribution of all patients that were involved in this study, by age and gender, whose data was extracted from NCD patient mastercards from 2020 to 2022. As such, a particular patient either had a non-communicable disease or a comorbidity of a non-communicable and HIV/AIDS. The average age of these patients was 30.9 years (SD = 6.02) and 68.3% of the patients were female. The young adult age group (19 – 39 years) being the subset of interest comprised of 10.6% females and 4.9% males of the total patients in the study.

**Table 4:** Gender versus age group distribution of cases.

| Age group | Gender | Freq (Perc) | CI |
| --- | --- | --- | --- |
| 0 - 18 | Female | 492 (2.77) | (2.02, 3.54) |
|  | Male | 507 (2.86) | (2.10, 3.62) |
| 19 - 39 | Female | 1887 (10.6) | (9.88, 11.4) |
|  | Male | 876 (4.94) | (4.18, 5.70) |
| 40 or more | Female | 9645 (54.4) | (53.6, 55.1) |
|  | Male | 4334 (24.4) | (23.7, 25.2) |

Table 5 shows the study demographics; gender, region, classification, and designation with respect to whether a patient had a non-communicable disease only or NCD-HIV/AIDS comorbidity specifically for young adults.

**Table 5:** Study demographics.

| Variable | Category | NCD Only |  | NCD-HIV/AIDS |  |
| --- | --- | --- | --- | --- | --- |
|  |  | Freq (Perc) | CI | Freq (Perc) | CI |
| Gender | Female | 1677 (60.7) | (58.8, 62.6) | 210 (7.60) | (5.72, 9.50) |
|  | Male | 836 (30.3) | (28.4, 32.2) | 40 (1.45) | (0.00, 3.34) |
| Region | North | 322 (11.7) | (9.70, 13.6) | 6 (0.22) | (0.00, 2.19) |
|  | Central | 808 (29.2) | (27.3, 31.2) | 10 (0.36) | (0.00, 2.34) |
|  | South | 1383 (50.1) | (48.1, 52.0) | 234 (8.47) | (6.51, 10.4) |
| Classification | City | 1143 (41.4) | (39.4, 43.4) | 97 (3.51) | (1.56, 5.53) |
|  | District | 1370 (49.6) | (47.6, 51.6) | 153 (5.54) | (3.58, 7.56) |
| Designation | Border | 260 (9.41) | (7.60, 11.3) | 4 (0.15) | (0.00, 1.99) |
|  | Inland | 1708 (61.8) | (60.0, 63.7) | 240 (8.69) | (6.88, 10.5) |
|  | Lakeshore | 545 (19.7) | (17.9, 21.6) | 6 (0.22) | (0.00, 2.06) |

The majority of the young adults’ females had a NCD only (60.7%) and 7.6% had a NCD-HIV/AIDS comorbidity. The southern region comprised of about 50% and 8.5% of NCD only and NCD-HIV/AIDS patients, respectively. There more NCD only (49.6%) and NCD-HIV/AIDS (5.5%) patients emanating from the district compared to cities where NCD only were 41.4% and NCD-HIV/AIDS were 3.5%. Inland locations also comprised of most of the NCD only and NCD-HIV/AIDS patients with 61.8% and 8.7% respectively followed lakeshore then border locations.

Table 6 Shows The progression of specific NCDs and NCD-HIV/AIDS cases through the three years.

**Table 6:** Summary of the case proportions by year.

| Type of Affliction | Year |  |  |
| --- | --- | --- | --- |
|  | 2020 | 2021 | 2022 |
|  | Freq (Perc) | Freq (Perc) | Freq (Perc) |
| Hypertension | 219 (7.92) | 259 (9.37) | 493 (17.8) |
| Asthma | 50 (1.81) | 124 (4.49) | 293 (10.6) |
| Diabetes | 113 (4.09) | 88 (3.18) | 143 (5.18) |
| Hypertension-Diabetes | 25 (0.90) | 35 (1.27) | 33 (1.19) |
| Epilepsy | 58 (2.10) | 135 (4.89) | 191 (6.91) |
| Other NCD | 53 (1.91) | 92 (3.33) | 109 (3.94) |
| Hypertension-HIV/AIDS | 0 (0.00) | 17 (0.62) | 63 (2.28) |
| Asthma-HIV/AIDS | 3 (0.11) | 2 (0.07) | 11 (0.40) |
| Diabetes-HIV/AIDS | 2 (0.07) | 3 (0.11) | 2 (0.07) |
| Hypertension-Diabetes-HIV/AIDS | 1 (0.04) | 1 (0.04) | 2 (0.07) |
| Epilepsy-HIV/AIDS | 4 (0.14) | 7 (0.25) | 8 (0.29) |
| Other NCD-HIV/AIDS | 5 (0.18) | 67 (2.42) | 52 (1.88) |

Hypertension had a year-by-year increase among the young Adults between 2020 and 2022. Hypertension-HIV/AIDS had a similar progression; diabetes hypertension-diabetes and diabetes-HIV/AIDS fluctuated through the years whereas the other types of affliction had year by year increase but rather with lower numbers than hypertension and hypertension-HIV/AIDS.

The comorbidity of a specific NCD and HIV/AIDS was present in about 9% of the patients. Of those, there was 2.9% comorbidity patients who had hypertension and HIV/AIDS (**Fig 2**).

**Fig 2.**
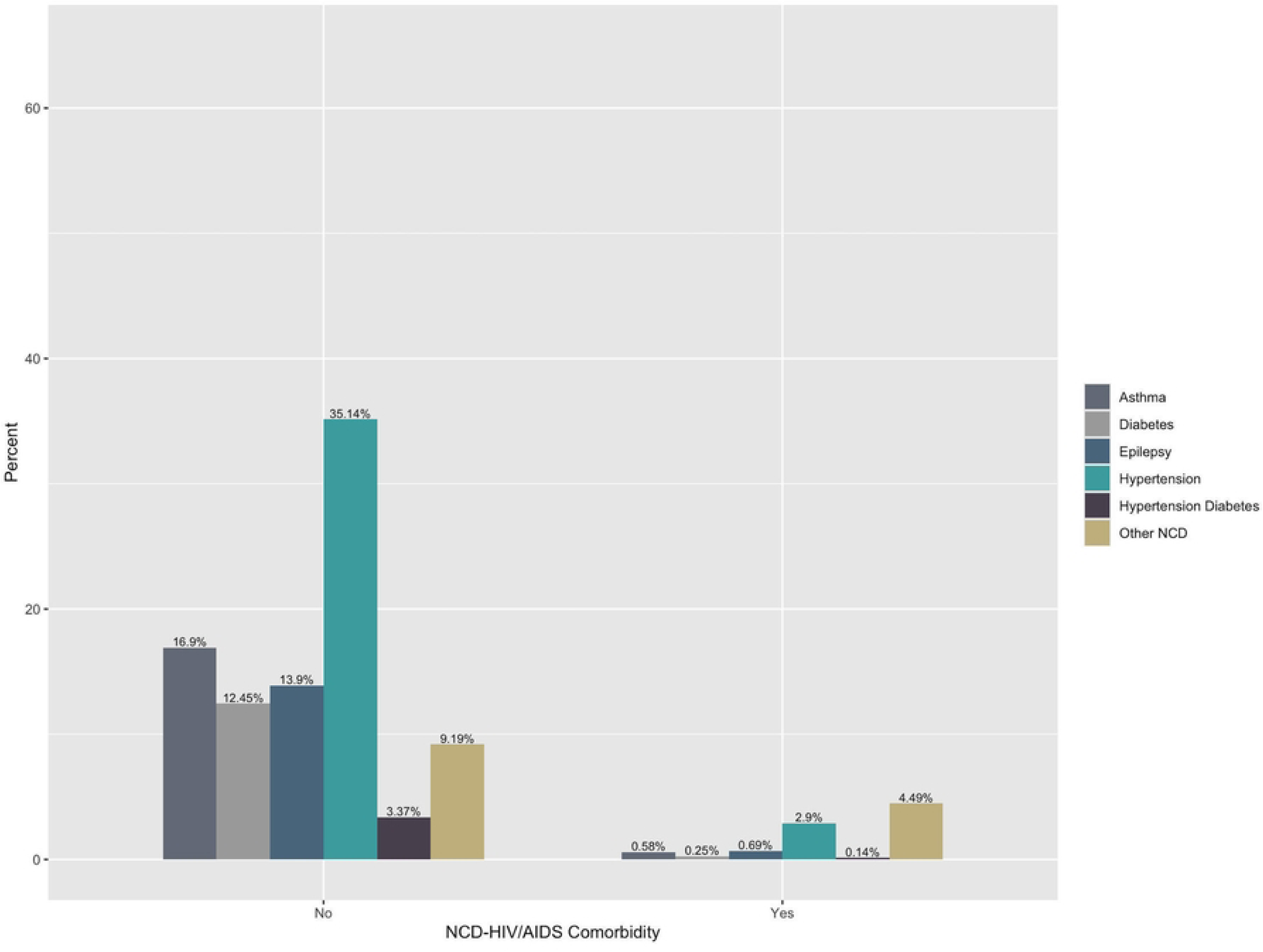
Specific NCD and NCD-HIV/AIDS comorbidity association.

There was also sufficient evidence to indicate that a relationship exists between a specific NCD and the presence HIV/AIDS (Chi-square = 314.44, p-value < 0.0001).

Table 7 shows Chi-square tests of association that were conducted between the outcome features, and the predictor features to determine if the predictors had a significant influence on the outcome.

**Table 7:**
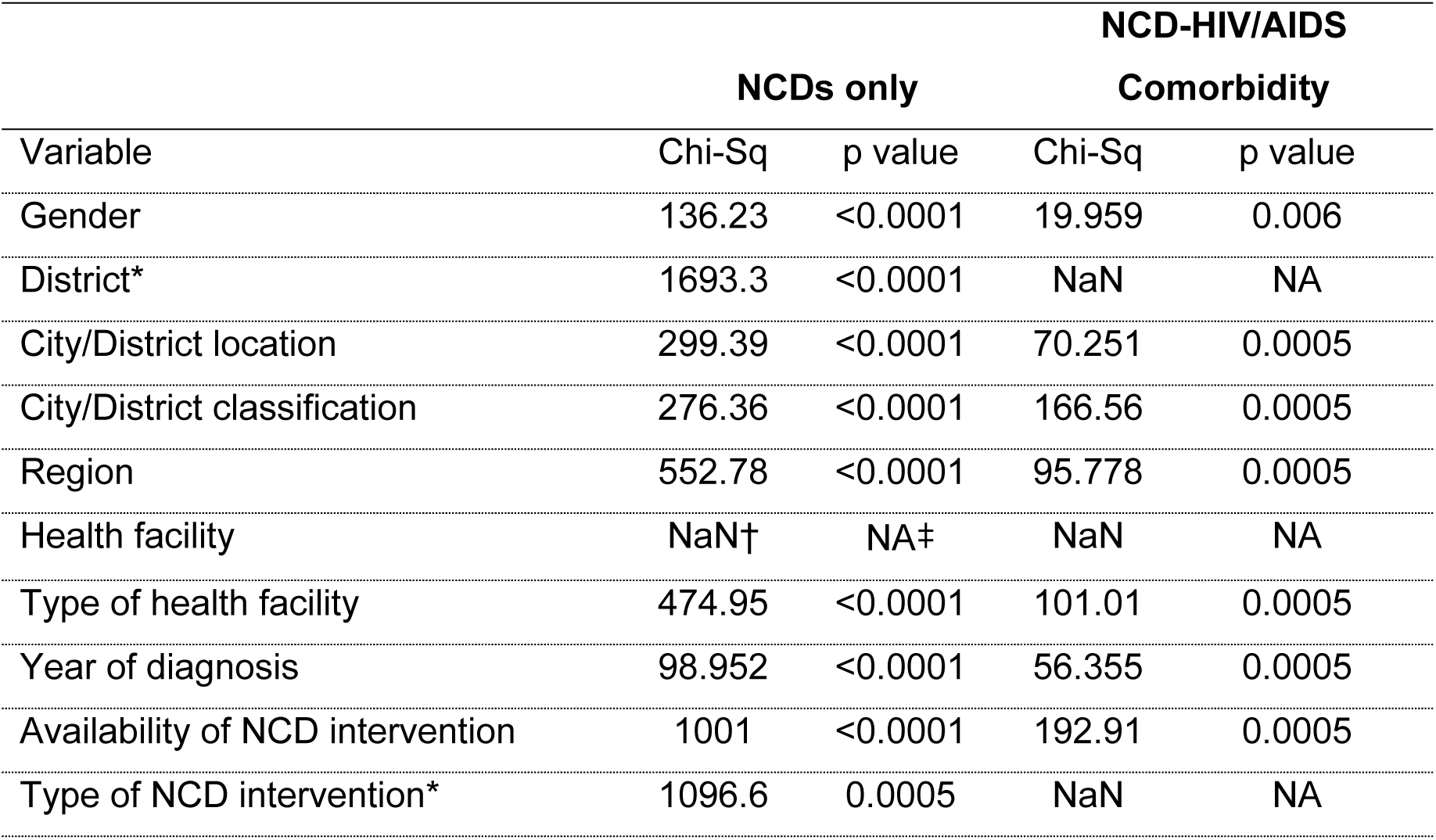
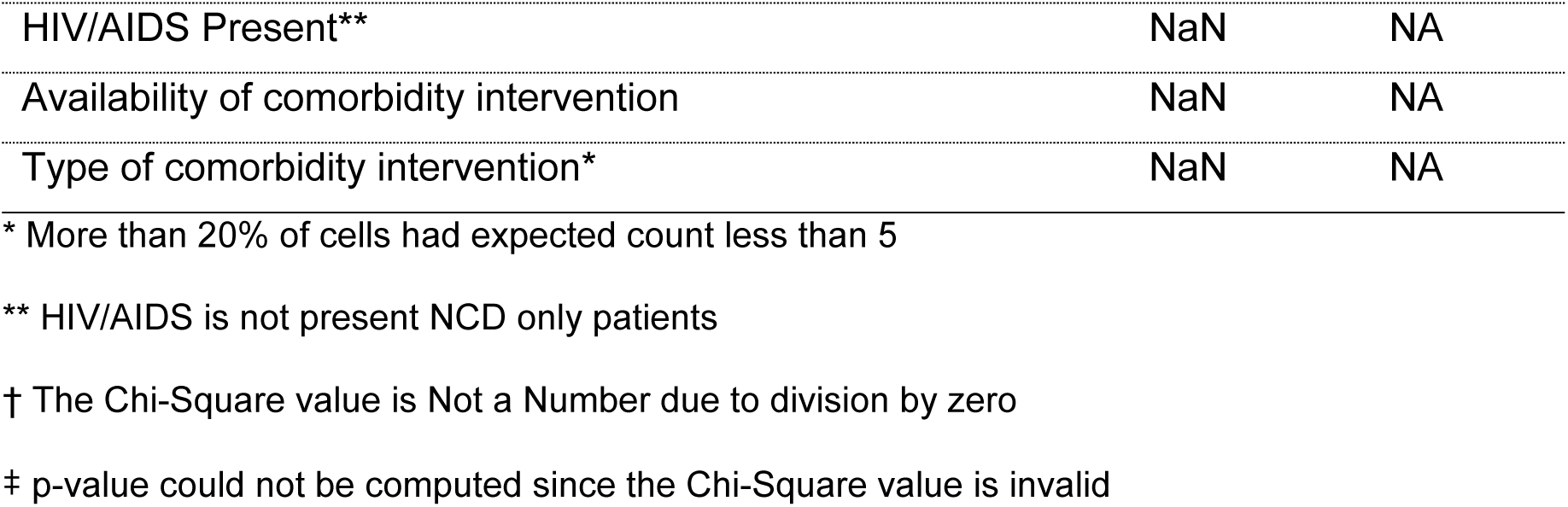
Associations between the response having a comorbidity against the predictors for young adult patients.

Gender had a significant association with specific NCDs (Chi-Sq. = 136.23 p < 0.0001) and specific NCD-HIV/AIDS comorbidity (Chi-Sq. = 19.595 p = 0.006). Table 7 shows that city/district location, city/district designation, region, type of health facility, year of diagnosis and availability of NCD intervention had a significant association both NCD and NCD-HIV/AIDS comorbidity outcomes. The type of NCD intervention was significantly associated with NCDs (Chi-Sq. = 1096.6 p = 0.005). The other predictors features: age group, residence, district, and health facility were not significantly associated with both NCD and NCD-HIV/AIDS comorbidity. The presence of HIV/AIDS, availability of comorbidity intervention and type of comorbidity intervention were specific features to NCD-HIV/AIDS comorbidity but were not significantly associated.

The area under the receiver operating characteristic curve (AUC-ROC) is used to evaluate the overall goodness of the models on NCDs and NCD-HIV/AIDS. This is a robust metric compared to the other metrics which only assess a single threshold. Considering the imbalance in our data, the hypertension and hypertension-HIV/AIDS cases were higher than the other classes as such the AUC-ROC was the best overall metric of choice due to its reliability. Fig 3 shows the NCD Only and NCDHIV/AIDS Comorbidity Cases Model ROC.

**Fig 3.**
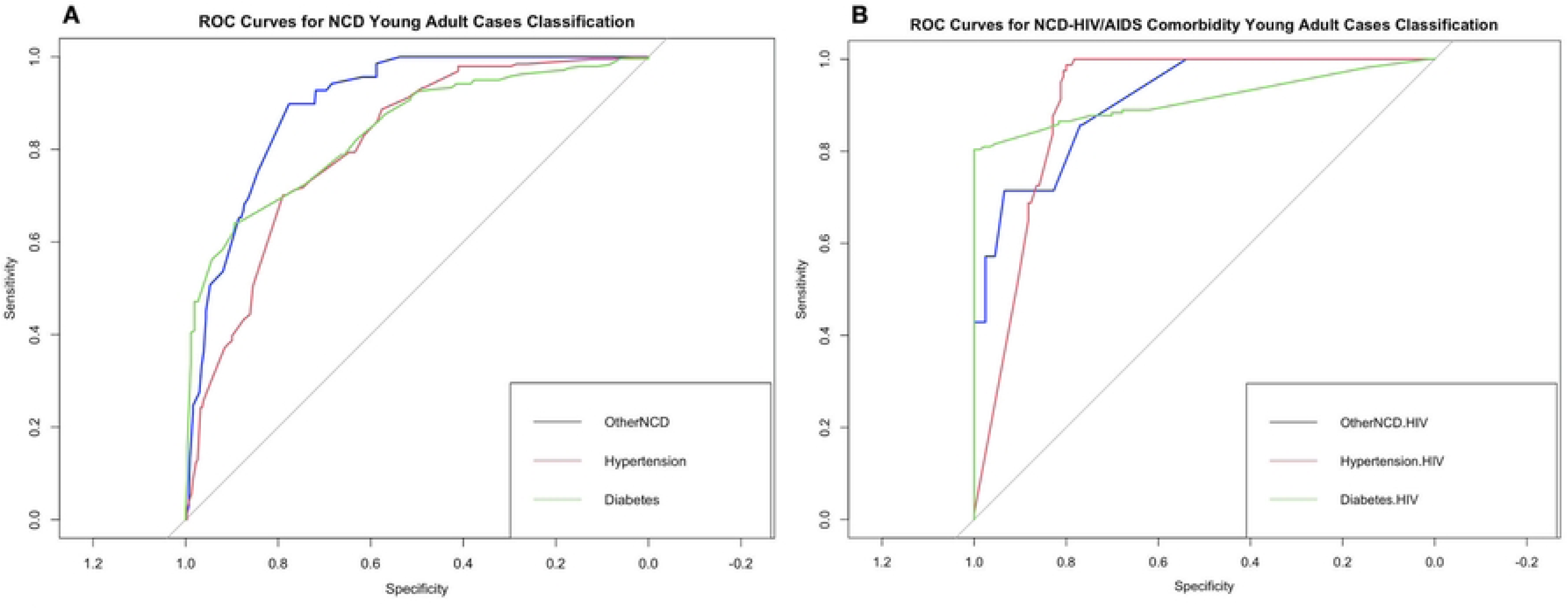
ROC Curves NCD only cases and NCD-HIV/AIDS comorbidity cases.

Fig 3 shows a plot for NCDs where the curve for each NCD is close to 1 indicating particularly good performance. The model was able to distinguish among the classes and classify the patients accordingly using given input features. The NCD-HIV/AIDS comorbidity model indicates better performance with curves for each outcome value even closes to 1 as shown in Table 9.

The confusion matrices for NCD only and NCD-HIV/AIDS comorbidity show the target and prediction classification in class-wise distribution. The machine learning model predictive performance is further evaluated using the confusion matrix (Fig 4).

**Fig 4.**
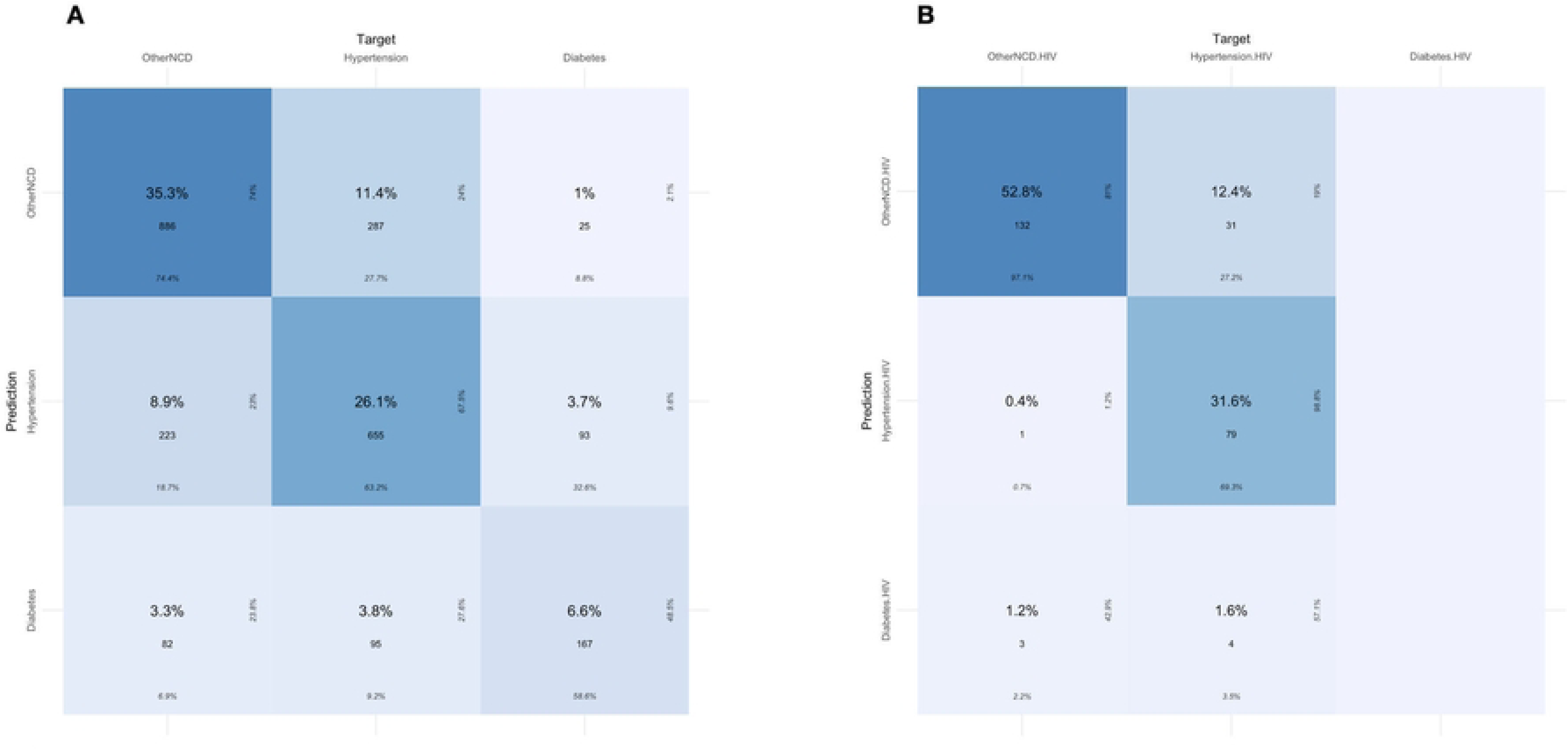
NCD only and NCD-HIV/AIDS Confusion Matrix.

Table 8 displays the global accuracy of the NCD only model and NCD-HIV/AIDS model were 68% and 84%, respectively. Other global metrics relating to the models were the AUCs (85% And 91%) complementing the ROC curves, Precision (48% and 45%) which considers that sample of the predicted class to actually belong to the positive class of the model, recall (49% and NaN) which is the number of correctly predicted samples in the positive class out of all the sample that belong in the positive class, and F1 - score (48% and NaN) for the positive classes whose values are the harmonic mean of the recall and precision scores for the NCD only and NCD-HIV/AIDS comorbidity models respectively.

**Table 8:** Metrics for NCD only and NCD-HIV/AIDS Model.

| Model | Accuracy (%) | AUC | Precision | Recall | F1-score |
| --- | --- | --- | --- | --- | --- |
| NCD Model | 0.680 | 0.850 | 0.475 | 0.490 | 0.481 |
| NCD_HIV/AIDS Model | 0.844 | 0.910 | 0.449 | NaN<sup>a</sup> | NaN<sup>a</sup> |

<sup>a</sup>Recall and F1-scores for Diabetes-HIV/AIDS were reported as NaN because there were no true instances for this class in the test data.

The confusion matrix (**Error! Reference source not found.**) further provided the ability to perform class-wise metrics for the models as shown in Table 9.

**Table 9:** Metrics for NCD only and NCD-HIV/AIDS comorbidity individual classes.

| Type of NCD and NCD-HIV/AIDS Comorbidity | Accuracy (%) | AUC | Precision | Recall | F1-score |
| --- | --- | --- | --- | --- | --- |
| Hypertension | 0.6320 | 0.8033 | 0.675 | 0.632 | 0.652 |
| Diabetes | 0.5841 | 0.8470 | 0.4855 | 0.586 | 0.531 |
| Other NCD | 0.7432 | 0.9004 | 0.740 | 0.744 | 0.742 |
| Hypertension-HIVAIDS | 0.6930 | 0.9079 | 0.810 | 0.971 | 0.883 |
| Diabetes-HIV/AIDS | 0.0000 | 0.9141 | 0 | NaN<sup>b</sup> | NaN<sup>b</sup> |
| Other NCD-HIV/AIDS | 0.9706 | 0.9068 | 0.988 | 0.693 | 0.814 |

<sup>b</sup>Recall and F1-scores for Diabetes-HIV/AIDS were reported as NaN because there were no true instances for this class in the test data.

The NCD only model was able to correctly classify 63.2% of the hypertension patients in that class. The precision, recall and F1-score were all above 60% for the hypertension class which implied that the model did fairly well in identifying and classify hypertension cases. The hypertension-HIV/AIDS class was accurately classified about 69% of the time. The other metrics were above 80% indicating that the model had remarkably high chances of classifying patients to this category. The diabetes and diabetes-HIV/AIDS classes had the least accuracies of 58% and 0% as well as the least precision, recall and F1-scores compared to other classes in their respective models.

The models were able to determine which feature were of utmost importance in classifying NCD only and NCD-HIV/AIDS patients. These features were categorized in order from the highest to lowest contribution in the models and presented in a feature importance bar graph (**Fig 5**).

**Fig 5:**
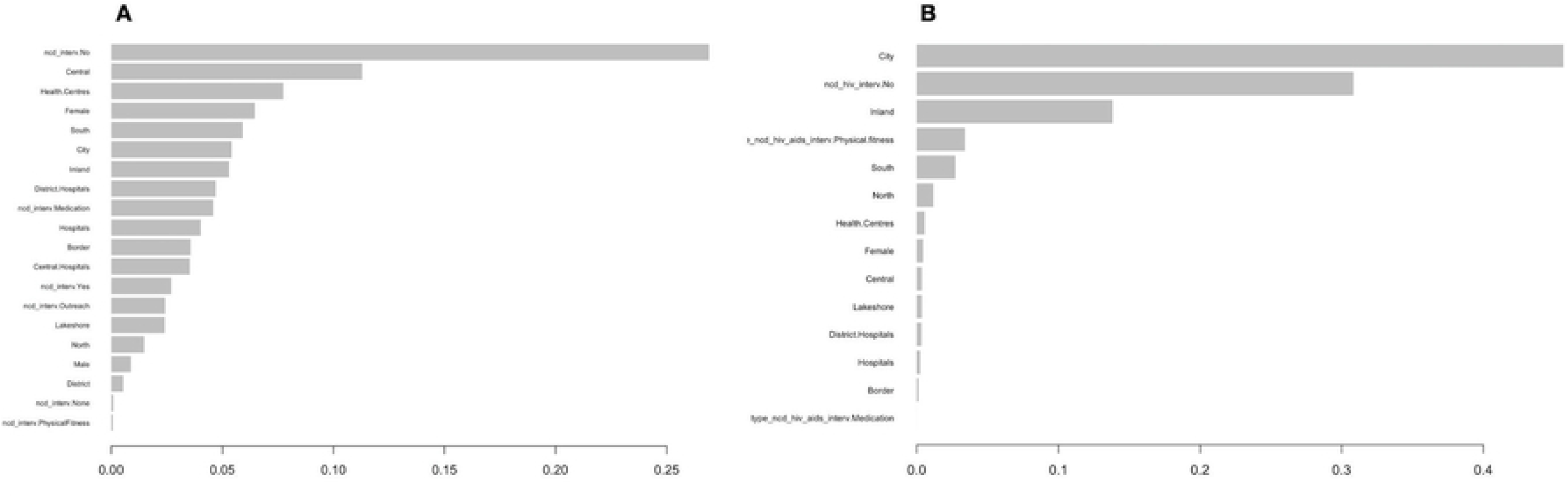
NCD-Only cases and NCD-HIV/AIDS comorbidity cases feature importance.

Fig 5A indicates the NCD only model had a patient having no intervention ranked highest (26.8%) in determinants of an individual having a hypertension, diabetes and other NCDs. An individual residing in central region of Malawi (10.5%), followed by being diagnosed at health centre health facility (8.1%), being female (6.7%), staying in the southern region (5.4%), staying in a city (5.6%) and the location being inland 5.4%), were the features significantly influencing having hypertension, diabetes and other NCDs. Low ranking features in the classification of NCDs were executing actual interventions, such as physical fitness (0.03%), no specific intervention (0.16%), residing in a district (0.51%) and being male (0.53%) in ascending order.

Fig 5B shows the features of the NCD-HIV/AIDS comorbidity model by order of importance in terms of their contribution to the model from high to low. The most contribution was from the feature living in a city (45.6%) and preceded by not having any interventions (30.8%) for the hypertension-HIV/AIDS, diabetes-HIV/AIDS, and other NCD-HIV/AIDS. A follow up substantial contribution to determining the specific NCD-HIV/AIDS comorbidities was when an individual dwelt in an inland location (13.8%). The feature with the least contribution to determining the comorbidities was an actual intervention where an individual is taking medication (0.004%). The location classification (border and lakeshore), type of health facility (hospitals, districts hospital, and health Centre), and the female gender had considerable contribution to the classification of the various NCD-HIV/AIDS comorbidity.

## Discussion

The study aimed to develop models for predicting NCDs and NCD-HIV/AIDS comorbidity given a patients’ socio-demographic characteristics. Extreme Gradient Boosting classification model was used to develop the NCD model and the NCD-HIV/AIDS model; classifying and predicting hypertension, diabetes, other NCD and hypertension-HIV/AIDS, diabetes-HIV/AIDS, other NCD-HIV/AIDS respectively. The models would enable easier and early detection of non-communicable diseases and NCD-HIV/AIDS among individuals.

Non communicable diseases, HIV/AIDS and NCD-HIV/AIDS affect individuals of all age groups [10,20]. This study specifically concentrated on NCD and NCD-HIV/AIDS patients aged 19 to 39 years, termed young adults, but however the number of patients with these ailments were increasing as the population got older [19]. Non-communicable diseases have had a reputation of being diseases among older People until recently [19]. This also concurs with the fact that People living with HIV are now living longer due to Antiretroviral Treatment which in turn leaves them susceptible to non-communicable diseases [22,24]. Special attention has been given to young adults in this study who are also affected by NCDs and HIV/AIDS [26,27]. These individuals are in their prime of productivity hence being the major contributor to the economy [26]. If young adults are afflicted by non-communicable diseases or NCD-HIV/AIDS, the implication is loss in productivity due to days off from sickness which affects the economy of the nation [28,29,43].

The female gender was the mostly affected with NCDs and NCD-HIV/AIDS comorbidity compared to males [11,49,68,69]. According to studies on a global scale and within region trends show that women are the mostly affected but however, women tend to report illnesses more than males [70]. Thus, the small numbers in males could be attributed to lack of reporting since other studies have reported disease burden in males [71]. A different view defines males as being more active which would lead to lower NCD cases, but it does not explain lower HIV/AIDS and NCD-HIV/AIDS cases [19,72].

The southern part of Malawi comprises of two cities which would account for more cases as NCDs and HIV/AIDS have high risk factors in cities [16,29]. The largest city in the Southern region, Blantyre, is a commercial City which implies rapid economic activities, growth in urbanisation and industrialisation [14,18]. These are all proponents of increase in NCD and NCD-HIV/AIDS cases as they drive risk factors for both diseases. Another notable location is Neno district which also drove the number of cases in the southern region higher providing a variation in the geographical location with varied risk factors [16]. Neno is one of the districts with a robust data collection system especially on NCD data courtesy of Partners Health (PIH) in collaboration with the Ministry of Health (MOH) [73]. The partnership aimed at boosting healthcare services with specific interest on non-communicable diseases and HIV/AIDS [74,75]. As such, all Health facilities in Neno are on the NCD Mastercard program and data is stored digitally.

Although districts seemed to have more patients compared to cities, it should be highlighted that there were 4 cities compared to 7 districts which implies that more patients were in cities. Districts having more patients may also not be considered an accurate picture due to a number of reasons; Neno is among districts and has all health facilities providing patient data and the major cities only have a fraction of health facilities on the NCD Mastercard program hence the cases are definitely under-reported [49].

In general, the non-communicable diseases and NCD-HIV/AIDS comorbidity are on an increasing trajectory. The specific top 5 NCDs and NCD-HIV/AIDS showed that hypertension was the leading NCD among the young adults. Globally hypertension is the prominent NCD as evidenced by the number of studies conducted on hypertension [10,15,21,76]. The trend is similar in the sub-Saharan region where hypertension is the leading affliction [14,21]. The other NCDs and their comorbidity with HIV/AIDS are equally important but more attention should be given to hypertension in designing and planning intervention.

The existence of a significant association between HIV/AIDS present in an individual or not and a specific non-communicable disease substantiates the coexistence of comorbidity cases among the population in Malawi [21,49,77]. Non-communicable diseases are disastrous in their individual existence in an individual and so is HIV/AIDS [10–12]. This speaks volumes to the gravity of the situation when an individual has the comorbidity of NCD and HIV/AIDS especially among young adults who are expected to be drivers of the economy.

Further tests were conducted to ascertain the association between specific NCDs and NCD-HIV/AIDS comorbidity against the socio-demographic features [78,79]. Gender, city/district location, city/district designation, region, type of health facility, and availability of NCD intervention were important features to determine a particular NCDs and NCD-HIV/AIDS comorbidity. Hence these features were used in the classification models.

Extreme Gradient Boosting (XGBoost) was used to conduct the classification of NCDs and NCD-HIV/AIDS due to its superiority and versatility [80]. XGBoost is a machine learning algorithm developed by Chen based on the gradient boosting algorithm [47].

Machine learning (ML) approaches in analysing data have quickly evolved into different fields of study including health and as applied in this study [81]. Machine learning algorithms have demonstrated their ability to give accurate and trustworthy predictions given past experiences [82]. Furthermore, ML algorithms are more flexible and efficient, easily scalable and deployable for use, which makes them more versatile to implement [82]. In comparison to conventional statistics, ML approaches do not have many strict assumptions that would render results insignificant if not satisfied [81]. In addition, ML approaches tend to produce better results unlike convention statistics thereby having better predictions [83].

Despite ML algorithms seemingly having an upper hand, conventional statistics or methods such as linear regression and multilevel models have provided tangible results as well [84,85]. Hence it is reasonable to consider conventional statistical methods and machine learning methods as complimentary tools to achieve different tasks [81]. For instance, Chi-Square test, Principal Component Analysis, Linear Discriminant Analysis, t-test and multicollinearity have been used to identify significant features and classify features applicable to certain problems and enable further analysis through machine learning using the identified features [81,86,87].

For the classification exercise in this study, logistic regression, multinomial logistic regression and its variants where considered. However, the model assumptions required to validate the results such as significance of the features and linearity of the Logit where violated. Therefore, conclusions could not be drawn from the models with such results. We further explored a deep learning technique, neural networks for this classification exercise which performed better than the conventional statistics despite lack of consideration for the imbalance in the data since most of the records were hypertension cases. It is important to recognise that the data imbalance is inherent to the record since hypertension was the most common non-communicable disease.

There are many machine learning algorithms that could be used in classifying NCDs and NCD–HIV/AIDS. These algorithms have been employed in various health studies in classifying different health problems [80,88–90]. Diverse applications of machine learning algorithms including decision trees, logistic regression, naive bayes, K–NN, Support Vector Machine (SVM), random forest, K–means, gradient boosting algorithms, lightGBM, CatBoost, GBM and XGBoost have also been applied in studying noncommunicable diseases [84,91–95].

The area studied ranged from classification of different stages of non-communicable diseases [93–96]. Extreme gradient boosting algorithm has been instrumental in modelling NCDs involving classification and prediction of risk levels of particular NCDs such as different levels of diabetes, different types of cardiovascular diseases, levels of disorder in anxiety, depression and stress, and severity of effects of treatment in different cancer patients [93–96]. XGboost produced better classification results and predictions compared to other ML models used [96,97]. Therefore, XGBoost had better predictive performance that was evidenced through the various performance evaluation metrics [93,94,98].

A study similar to this study considered classifying the following NCDs; hypertension, stroke, cardiovascular diseases, and diabetes using various machine learning techniques. The main objective was to classify multi-NCDs and in this study the Random k-labelsets machine learning technique was able to achieve the highest accuracy [99]. In contrast, our study develops two models classifying diabetes, hypertension and other NCD as well as diabetes–HIV/AIDS, hypertension–HIV/AIDS and other NCD–HIV/AIDS using XGBoost algorithm respectively. XGBoost is a more efficient and scalable machine learning algorithm [80]. It was chosen for its reputation from Kaggle competitions and other studies that employed it solving various forms of classification problems [93].

Machine learning can be a tool that promotes early prediction and detection of diseases thereby enabling development of management and intervention strategies [95]. Extreme Gradient Boosting was used for early detection and prevention in many studies including a study by Davagdorj where classification and predictions were made on the risks of a smoke induced NCDs [98]. As such, we applied the XGBoost algorithm to classify and predict non-communicable diseases and NCD–HIV/AIDS comorbidity for early detection and implementation of management and intervention strategies.

Features enable the XGBoost algorithm model the problem, correctly classifying and making predictions of the outcomes [86,97,98]. A vast number of features would affect an individual having an NCD or NCD-HIV/AIDS comorbidity. This study involved modelling NCD and NCD-HIV/AIDS cases using socio-demographic features. The Chi-Square test was used to determine features that had significant influence, by associating with either a specific NCD or NCD-HIV/AIDS, to be included in the future set for each model [97,98]. The XGBoost algorithm then mapped, in descending order, feature importance according to how much contribution was provided by each feature to the classification and prediction [86,97].

The model metrics enabled evaluation of different NCD and NCD-HIV/AIDS comorbidity models through an iterative process until suitable models were obtained [60–62]. The chosen models had the required predictive power to determine the particular NCD or NCD-HIV/AIDS comorbidity an individual will be afflicted with given a set socio-demographic characteristic [78].

The results established a set of features that may be used to determine an individuals’ risk factors to having a specific NCD and NCD-HIV/AIDS comorbidity. The socio-demographic features enabled the classification and prediction of NCD and NCD-HIV/AIDS cases. The models ranked the different predictor features in order of importance depending on how much each feature contributed to the classification of the outcome feature [62,67]. The NCD model has lack of intervention as having the most contribution in determining whether an individual had hypertension, diabetes or other NCD while the NCD-HIV/AIDS model had ranked no intervention as the second in classifying hypertension-HIV/AIDS, diabetes-HIV/AIDS, or other NCD-HIV/AIDS. The knowledge of NCDs and their risk factors through sensitization provides individuals with a platform to control modifiable risk factors to reduce the likelihood of having a non-communicable disease [71]. The gender female was also on influence in classifying NCD cases and this fact resonates with the number of cases reported in this study and other studies [11,68,69]. However, the NCD-HIV/AIDS did not classify female gender highly. Location was also one of the crucial factors for both models in the classification since these locations have growing economic activities, increased industrialisation and urbanisation which are all drivers of NCD and HIV/AIDS risk factors [13,14]. The factors determined by the model would enable reduction of time in diagnosis of NCD and NCD-HIV/AIDS cases. Furthermore, depending on an individuals’ features, a set of risk factors would be determined and enable implementation of practical interventions to manage and prevent case occurrence. Quick and early diagnoses would be plausible since the models can be deployed for use at different health facilities. Model deployment involves saving the model, loading the model for prediction and use of an Application Programming interface (API) to run the model at a point of service. The API, usually a web service or an app, would be used to input data on patients’ social demographic factors to either the NCD only or NCD-HIV/AIDS trained models which would return a predicted NCD or NCD-HIV/AIDS ailment. Therefore, health personnel would have a head start in diagnosis by first considering the specified NCD or NCD-HIV/AIDS case classified and conduct tests based on the model output.

## Conclusions

The study sought to develop models to aid in predicting NCD and NCD-HIV/AIDS comorbidity cases using a set of socio-demographic factors. Extreme Gradient Boosting for classification was the chosen methodology in classifying NCD and NCD-HIV/AIDS comorbidity using the influence of factors such as gender, location and availability of intervention. Its ability to tune hyper-parameters proved its strength in the ability to provide sufficient models for the given data. The models provided tangible results for modelling non-communicable diseases and the comorbidity of NCD and HIV/AIDS. Given a particular set of features, the model was able to distinguish a hypertension patient to a diabetes patient, other NCD patients and vice versa. In a similar manner, the model also distinguished a hypertension-HIV/AIDS patient to a diabetes-HIV/AIDS patient, other NCD-HIV/AIDS patient and vice versa.

The XGBoost models were able to accurately predict an ailment a patient would be afflicted with, considering a set of socio-demographic characteristics about the individual. The models are of particular significance because they can be deployed using an API and used to make predictions on which ailment an individual may have and reduce doctors’ time in conducting so many tests to diagnose for an ailment. This reduces the work required at micro decision level.

Further from predictions, the model indicated socio-demographic factors that highly influenced proneness to a particular NCD and NCD-HIV/AIDS which can be used to design data driven intervention and influence macro decisions to prevent an increase in the number of cases. This would improve the progress towards achieving Sustainable Development Goal 3.3 and 3.4.

## Strengths and Limitations

The strength of this study is that it delves into the use of machine learning approaches, leveraging their versatility and high predictive power compared to traditional statistical methods that are less versatile. Furthermore, the study encompasses 4 cities and 7 districts in Malawi providing a wide socio-demographic canvas thereby boosting the application of the predictions to cities and districts with similar characteristics.

In terms of limitations, the data collection process did not utilise artificial intelligence techniques which would have been easier and faster. However, the required data to create a database for modelling the NCD and NCD-HIV/AIDS comorbidity as well as specific NCDs and their comorbidity with HIV/AIDS was obtained.

The dataset was imbalanced with majority of the cases being hypertension and hypotension-HIV/AIDS cases which usually causes problems for classification models. Despite this being a limitation for classification models, it shows an inherent characteristic in the population that hypertension is a big problem that needs to be considered with utmost importance. However, XGBoost is a versatile model with so many parameters that aid in managing imbalanced datasets during the classification process and produce reliable results.

Another limitation of the study was lack of incorporation of covid19 interaction in the models. The data from NCD Mastercards does not have any covid19 features /variables that would enable modelling with covid19 to determine its influence.

Social demographic features only were used to model NCDs and NCD-HIV/AIDS comorbidity. There are other features such as medical indicators which should play a pivotal to role in the classification and improved model predictive accuracy. Considering the magnitude of this study as well as the sources of data, medical indicators could not be obtained. As such, the addition of medical indicators is part of our future work as we continue to improve the models to classify and predict more NCDs and NCD-HIV/AIDS comorbidity.

## Author contributions

AK was responsible for data extraction; MK and AK conceived, designed and planned the study; AK analysed and interpreted the data and wrote the first draft of the manuscript; SM and AM interpreted the data and wrote the final draft of the manuscript together with MK and AK. All authors have critically reviewed and approved the final version of the manuscript.

## Data availability

Data for the study are confidential.

## Declarations

### Ethical approval and consent to participate

Ethical clearance for the study was obtained from the National Committee on Research in Social Sciences and Humanities (NCRSH), an ethics committee operating under the National Commission for Science and Technology (NCST). The protocol reference number is P.01/1/22/613.

## Acknowledgements

We sincerely thank the patients, the data clerks at the health facilities and District Health Office DHMTs for their support of this project. In particular, special thanks to the research assistants; Steven Kayipa, Madalitso Tambala, Emmanuel Guzani, Alfred Ephraim, Hanna Bello, Emily Chisale, Alick Doben, Wiseman James, Lazarus Bamusi and Mariam Muhajiri who were instrumental in data extraction.

